# Treatment Resistant Depression in electronic health records: Definitions Matter

**DOI:** 10.64898/2025.12.15.25342069

**Authors:** Matthew H Iveson, Emily L Ball, Chris Wai Hang Lo, Matúš Falis, Cathryn M Lewis, Heather C Whalley

## Abstract

**Background:** Many people with depression do not respond well to the first antidepressant prescribed. Treatment Resistant Depression (TRD) refers to depression which does not respond to multiple subsequent antidepressant treatments. Identifying TRD in routinely-collected health records is challenging due to limited response-related data. Previous studies have used definitions based on the number of antidepressant switches observed. However, these do not account for other features clinically indicative of treatment resistance, such as augmentation of antidepressants with lithium or antipsychotics and switches between antidepressant classes. This study examined definitions of TRD and their impact on the resulting sample across three cohorts.

**Methods:** Across the DataLoch, UK Biobank, and Generation Scotland cohorts, we identified cases of depression from primary and secondary care record codes and extracted antidepressant treatment patterns from dispensing/prescribing records (N = 51,283, N = 10,556, and N = 649 respectively). We examined 9 TRD definitions that varied by: the number of switches required (1+, 2+ or 3+ switches), augmentation and between-class switches. We contrasted sample size and characteristics between definitions, and examined factors associated with inclusion versus a reference definition of 2+ switches.

**Results:** The reference TRD definition included 10% of depression cases in the routine data collection, but substantially fewer cases (4%) in consented cohorts. More inclusive definitions that required fewer switches or included a between-class switch classified more individuals as TRD, but resulted in a proportionally older, more deprived sample with fewer depression-related health record codes, older age of depression onset, lower symptom severity, and greater use of first-line antidepressants. Requiring more switches (3+ switches) classified fewer individuals as TRD, but resulted in a proportionally younger sample, with more depression-related health record codes, younger age of depression onset, and greater use of antidepressants associated with later in the treatment line (e.g., Tricyclics). Definitions including augmentations resulted in a small increase in sample size without notable change in sample characteristics.

**Conclusions:** TRD is underrepresented in consented cohort studies. A definition of TRD that includes 2+ antidepressant switches or augmented antidepressant treatment as indicators balances sample size with depression severity, while incorporating features from real-world treatment journeys.

**Key summary:**

- Treatment Resistant Depression (TRD) is often identified in health records using number of switches in antidepressant treatment, but this misses other important indicators of treatment resistance
- We examined 9 TRD definitions across 3 cohorts, varying in the number of antidepressant switches, the inclusion of augmented treatment, and the inclusion of between-class switches
- A TRD definition that included 2+ switches or augmented treatment balanced sample size and severity

## Introduction

Many individuals treated for depression do not respond well to initial antidepressant treatment. Current guidance generally recommends commencing antidepressant treatment with a Selective Serotonin Reuptake Inhibitor (SSRI) and to consider switching to another antidepressant if there is no sufficient improvement after adequate time and dose (National Institute for Health and Care Excellence, 2022; Taylor et al., 2021). The failure of multiple subsequent treatments, termed Treatment Resistant Depression (TRD), occurs in around a third of patients (Souery et al., 2011; Trivedi et al., 2006) and can have severe consequences for health and wellbeing (Armbrecht et al., 2021; Jensen et al., 2022; Mann et al., 2021; McIntyre et al., 2019, 2020; Sussman et al., 2019). However, the lack of an agreed definition of TRD has been identified as a significant barrier for research, practice and drug development (McIntyre et al., 2023).

Although defined in clinical trials using validated scale scores (e.g., Montgomery-Asberg Depression Rating Scale), TRD is harder to define in studies using real-world data from electronic health records, as symptom information and scale scores are rarely available. Several working definitions have been proposed, with little consistency in the type or degree of criteria to be applied (McIntyre et al., 2023). Most commonly, studies have considered those with 2 or more switches in antidepressant treatments to be TRD-positive cases (Souery et al., 2007), with the caveat that treatments must have been of adequate dosage and length – generally taken to be a minimum treatment-effective dosage for 4-6 weeks (Blier, 2009; Kendrick et al., 2022). Notably, several clinical bodies (the European Medicines Agency (EMA), the National Institute for Health and Care Excellence (NICE) and the Food and Drug Administration (FDA)) have taken up this simple ‘two or more’ definition, with some small variation in detail. However, a definition including only switches does not reflect the complexities of real-world treatment journeys and does not take advantage of the opportunities of routinely-collected health records (Lo et al., 2025).

Firstly, not all switches in antidepressants are the same. Treatment guidelines recommend different classes of antidepressant at different points in the treatment line due to mechanistic differences, so between-class switches may be more informative than within-class switches. For example, non-response to one SSRI (e.g., Sertraline) may be expected if non-response was previously noted to another SSRI (e.g., Citalopram), given their similar mechanism and pharmacokinetic profile (Laux, 2021). A switch from an SSRI to another class of antidepressant may be more indicative of treatment resistance than an SSRI-SSRI switch. Furthermore, switching to other classes of antidepressant, particularly Tricyclics (TCAs) and Monoamine Oxidase Inhibitors (MAOIs), is only recommended much later in treatment algorithms after first-line treatments have failed (National Institute for Health and Care Excellence, 2022). Previous definitions do not distinguish between within- and between-class switches due to their similar remission rates in clinical trials (Fabbri et al., 2017).

Secondly, in later stages of treatment, guidelines often recommend the augmentation of antidepressant therapy with other psychotropic medication (e.g., lithium, antipsychotics) to enhance effectiveness even in the absence of psychotic symptoms. While between-class switches and augmentation can be readily identified in prescription records, they are not typically included in TRD definitions applied to health record data (e.g., the EMA) (Committee for Medicinal Products for Human Use, 2023). Further indicators, such as receipt of electroconvulsive therapy (ECT) have been considered in somewhat older models of TRD (Thase & Rush, 1997), but are not referenced in modern, more widely-adopted models.

Notably, including between-class switches and augmentations has the benefit of accounting for limitations in data availability. Most sources of prescribing data are recent, for example Scottish routine dispensing data is available from 2009, and do not capture the full journey of people initiating treatment prior to this time. If an individual has experienced two distinct antidepressant treatments before electronic health records start and two after, a strict ‘2 switch’ definition would not classify them as a TRD-positive case (only 1 switch is recorded, of the 3). However, a TRD definition that included between-class switches would cover such a case, and so may be better at capturing those (predominantly older) individuals further into their antidepressant treatment journey at data start.

In the present study, we applied the existing definition of TRD across three distinct cohorts, including two consented health cohort studies and an e-cohort created from population health records. In these three cohorts, we examined how changing the threshold for number of switches, the inclusion of augmentation, and the inclusion of between-class switches changes the size and characteristics of the resulting TRD-positive sample.

## Methods

### Sample

Initial analyses were conducted using a data extract obtained after approval through DataLoch (https://dataloch.org/data/about-the-data), a service that enables access to de-identified healthcare data from the South-East Scotland region. This service covers over 90% of primary care services and all secondary care services in Lothian, and includes over 900k individuals alive at data access. Data was accessed in September 2024.

We conducted similar analysis in two consented health cohort studies with extensive electronic health record linkage – UK Biobank (UKB) and Generation Scotland (GS). Cohort studies were chosen to provide a more in-depth examination of correlates of TRD, particularly associations with symptom severity. UKB includes over 500k participants recruited from across the UK between 2006 and 2010 (Bycroft et al., 2018), while GS includes over 22k individuals recruited from across Scotland around 2012 (Milbourn et al., 2024). Both cohorts obtained consent for linkage of survey and biological data with routinely-collected health records for research purposes.

To create the analytic samples for each of the three cohorts, we first selected all individuals with at least 2 different Major Depressive Disorder (MDD)-related diagnostic codes in GP or hospital admission records (for DataLoch: READ v2 codes in GP records and ICD-10 codes in Scottish Morbidity Records 01 and 04, 2014-2024; for UKB: READ v2 and v3 codes in GP records and ICD-9 and -10 in Hospital Episodes, 1981-2017 depending on region; for GS: READ v2 in GP records and ICD-9 and -10 in Scottish Morbidity Records 01 and 04, 1980-2022). This initial step was done to increase the likelihood that included individuals were receiving antidepressant treatment for their depression, rather than for other conditions (e.g., neuropathic pain). We then removed all individuals with a GP or hospital admission record indicating drug misuse, alcohol misuse, bipolar disorder or psychosis. The resulting sample consisted of 51,283 individuals from DataLoch, 19,641 individuals from UKB and 649 individuals from GS. A sample flow diagram is shown in Figure 1, and diagnostic code lists and a breakdown of excluded participants are provided in Supplementary Material.

**Figure 1.**
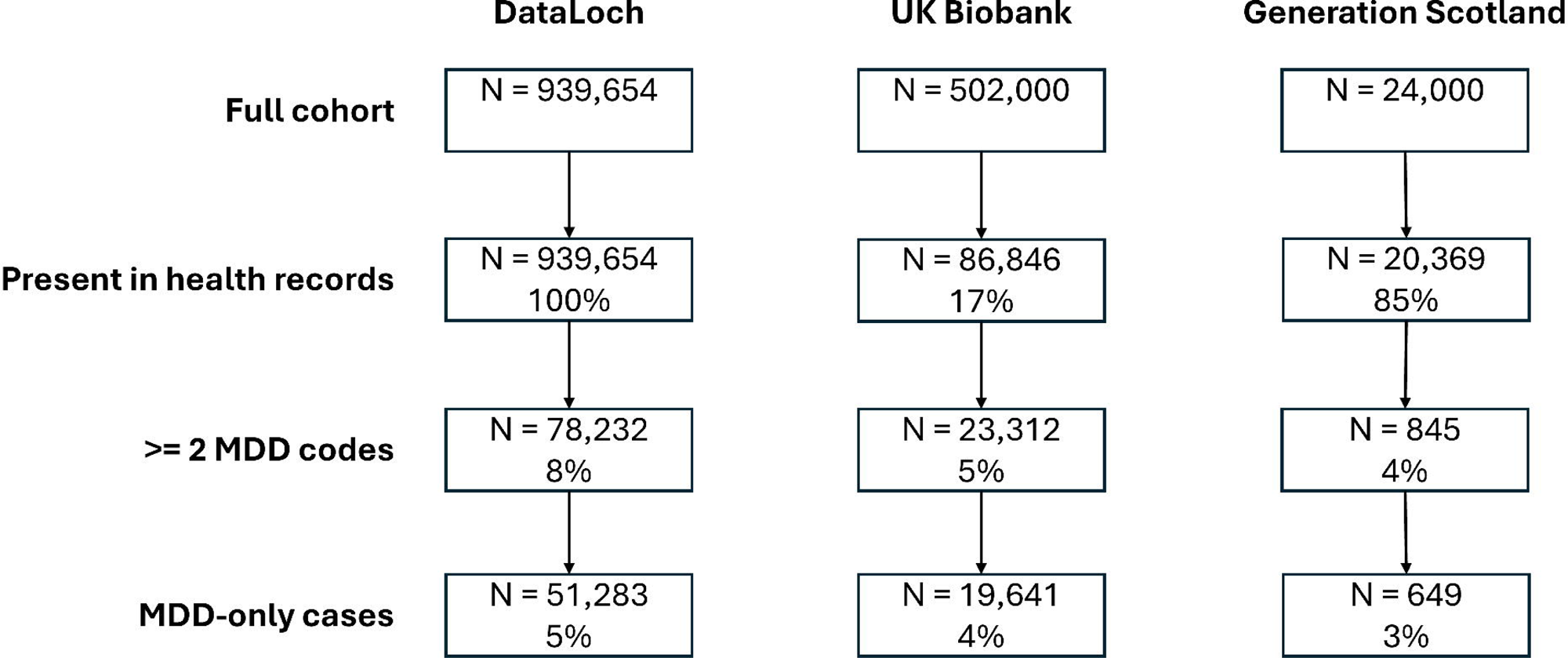
Sample flow diagram for DataLoch, GS and UKB showing selection based on being present in health records (hospital admissions or GP records), MDD diagnoses and other psychiatric diagnoses and percent of Full cohort.

### Measures

#### Prescribing episodes

Antidepressant prescribing data was extracted from linked prescribing records (for DataLoch: Prescribing Information System, 2015-2023; for UKB: Primary Care Prescribing, 1980-2017; for GS: Prescribing Information System, 2009-2021). All prescribing data was converted from event format to episode format by combining neighbouring (concurrent and within 26-weeks, see Fabbri et al., 2021) events of the same drug (approved item name), strength and formulation. That is, new prescribing episodes were identified with a change in approved item name or after a substantial gap in prescribing. Episode data was filtered to include only prescribing episodes lasting 6 weeks or longer, more conservative than NICE guidance on adequate treatment time for antidepressants (National Institute for Health and Care Excellence, 2022) to avoid changes due to side effects (Wigmore et al., 2020).

Antidepressant episodes were created by selecting episodes with BNF codes starting ‘0403’, excluding tryptophan.

#### Treatment Resistant Depression

TRD was defined using three features derived from prescribing data: switches, augmentation and between-class switches. Switches were defined as a new antidepressant episode (within 14 weeks of the previous episode) triggered by a change in the approved item name (i.e., changes in dosage, strength or formulation were not included), regardless of class. For example, an episode of fluoxetine that begins 10 weeks after an episode of citalopram would be identified as a switch. In line with previous work, only distinct switches were included (i.e., switching back to a previous antidepressant was not counted as an additional switch).

Augmentation was defined as an episode of a NICE-approved agent that overlapped an antidepressant prescription episode, including lithium and specific antipsychotics (see Supplementary Material for full list). Between-class switches were defined as a switch (see above) where the previous and current antidepressants came from different classes: Selective Serotonin Reuptake Inhibitors (SSRIs), Selective Noradrenaline Reuptake Inhibitors (SNRIs), Tetracyclics (TetCAs), Tricyclics (TriCAs), Monoamine Oxidase Inhibitors (MAOIs), and other. Classes were based on mechanistic similarity, not on BNF code sections (see Supplementary Material for a full list). In the earlier example, a change to fluoxetine from citalopram would be included as a switch, but not a between-class switch as both are SSRIs.

A total of nine definitions of TRD were created by varying the number of switches included (1-3), whether augmentation was included, and whether between-class switches were included. Any of the criteria could be met for inclusion. For example, individuals receiving augmented antidepressant treatment would be included in all of the definitions that included augmentation, regardless of how many switches they experienced. Similarly, individuals experiencing one SSRI-TCA switch would be included in all of the definitions that included between-class switches (as well as the 1+ switch definition).

#### Covariates

Covariates were chosen based on prior evidence of their association with depression and treatment resistance (e.g., Hölzel et al., 2011). Across the three cohorts, we extracted the number of distinct MDD-related diagnostic codes (ICD9/10 and READv2/3) across GP and hospital records as a proxy for severity. That is, more distinct diagnostic codes indicate more complex or changing depression as well as more healthcare utilisation. We also extracted the average length of antidepressant episodes (in months) across prescribing records, and the most recent antidepressant class from prescribing records.

In the DataLoch cohort, we also extracted age (in years) at the last GP or hospital record, the age (in years) at the first MDD-related diagnostic code across GP and hospital records, area-level relative socioeconomic deprivation (Scottish Index of Multiple Deprivation (SIMD) 2021 quintile; higher is less deprived), sex (from routine health records) and ethnicity (from routine health records; Asian, White, Other, Mixed, Not Stated).

In the UKB cohort, we extracted measures not available in routinely-collected health records (Supplementary Material). These included self-reported measures from various surveys and clinics throughout UKB follow-up: age of first depression diagnosis (in years), an indicator of family history of depression, an indicator of ever self-harmed, BMI, and highest educational qualification. Deprivation was measured using the Townsend Deprivation Index (Townsend et al., 2023), a relative indicator of material deprivation, which was reversed such that scores above 1 indicated lower area level deprivation. We also extracted measures of psychiatric symptom severity from standardised questionnaires, including Composite International Diagnostic Interview Short Form (CIDI-SF) total score and Patient Health Questionnaire 4 (PHQ4) total score (‘feeling down’, ‘little interest’, ‘feeling nervous’, and ‘worrying’ items from their worst depressive episode), as well as indicators of response to any antidepressants (including citalopram, fluoxetine, sertraline, paroxetine, amitriptyline, dosulepin, and other antidepressants) from a mental health survey (Kamp et al., 2025). We also extracted polygenic risk score for MDD, calculated using the Psychiatric Genomics Consortium MDD3 algorithm at a p-value threshold of <0.001 (Adams et al., 2025).

In the GS cohort, we extracted self-reported age of depression onset (in years), number of depressive episodes, and highest educational qualification. As with the DataLoch cohort, we also extracted SIMD quintile at study entry. We also extracted measures of psychological distress and mood symptom severity from standardised questionnaires, including the General Health Questionnaire (GHQ) and the Mood Disorder Questionnaire (MDQ), as well as neuroticism total score from personality questionnaires. As with the UKB cohort, we also extracted a polygenic risk score for MDD, calculated using the Psychiatric Genomics Consortium MDD3 algorithm at a p-value threshold of <0.001 (Adams et al., 2025).

### Statistical Analysis

For each of the three cohorts we applied each of the 9 TRD definitions. Sample size was calculated as the fraction of those in the definition versus the total analytic sample included in each definition (i.e., MDD patients exposed to antidepressants). Descriptive statistics were generated for each of the resulting samples. Against the reference definition of 2+ switches, the odds (ORs) of being included in a given definition associated with each available covariate individually were estimated using multinomial logistic regression models. For example, one model estimated the association between age and odds of being included in the 1+ switch definition (as well as all other definitions), versus the 2+ switch definition.

Supplementary analyses were run to investigate the impact of including receipt of ECT in the TRD definitions. Notably ECT is typically used as a treatment for severe depression when other treatments have been unsuccessful (National Institute for Health and Care Excellence, 2022), but requires additional data sources beyond prescribing and dispensing data. In this study, ECT data was only available for the DataLoch cohort, and was based on outpatient records.

Analysis was conducted in R (v4.4.2)(R Core Team, 2023) and RStudio (v2024.09.1)(RStudio Team, 2023). Ethical approval was obtained from the Edinburgh Medical School Research Ethics Committee at The University of Edinburgh (23-EMREC-035).

## Results

### Sample size

Of the MDD cases identified in each sample, around 70% had at least one adequate antidepressant episode of 6+ weeks (DataLoch = 70%, UKB = 50%, GS = 71%). The proportion of MDD cases included in each of the 9 TRD definitions are shown in Table 1. The reference TRD definition of 2+ switches captured 10% of the MDD sample in DataLoch, a population cohort, but only around 4% of the MDD sample in UKB and GS, both consented cohorts. Considering the number of switches, reducing the number of switches, resulted in a 1.6 – 5.0-times larger proportion included in the TRD sample, whereas increasing the number of switches, resulted in a moderately (0.25 – 0.6 times) smaller proportion included in the TRD sample. Adding augmentations resulted in a very small (around 1% of each cohort) increase in TRD sample size across definitions and across cohorts. Further adding 1+ between-class switches resulted in a substantial increase in sample size, particularly for the (least inclusive) 3+ switch definition (2.3 – 12.0 times). The increase from adding between-class switches was less pronounced for the 2+ switch definition (1.4 – 3.0 times). The sample sizes of the 1+ switch definitions were unsurprisingly unaffected by adding between-class switches, as these switches were already covered by the basic definition.

**Table 1.** The proportion of individuals included in each TRD definition, as a proportion of those in the analytic samples, for each cohort. Column headings describe the inclusion criteria for each definition, any of which can be met for inclusion.

|  | **1+ Switch** | **1+ Switch**<br>**Augmentation** | **1+ Switch**<br>**Augmentation**<br>**Class switch** | **2+ Switches**<br>**(REF)** | **2+ Switches**<br>**Augmentation** | **2+ Switches**<br>**Augmentation**<br>**Class switch** | **3+ Switches** | **3+ Switches**<br>**Augmentation** | **3+ Switches**<br>**Augmentation**<br>**Class switch** |
| --- | --- | --- | --- | --- | --- | --- | --- | --- | --- |
| **DataLoch**<br>N = 51,283 | 27% | 27% | 27% | 10% | 11% | 23% | 4% | 4% | 23% |
| **UK Biobank**<br>N = 19,641 | 4% | 4% | 4% | 3% | 3% | 4% | 1% | 1% | 3% |
| **Generation**<br>**Scotland**<br>N = 649 | 20% | 21% | 21% | 4% | 4% | 12% | 1% | 2% | 12% |

### Sample demographics

Sample demographics resulting from each of the TRD definitions, for each of the cohorts, is available in Supplementary Material. Briefly, individuals included in each definition were predominantly middle- and older-aged adults (median ages 44 – 64 years), predominantly female (64 – 77%), and predominantly white ethnicity (90 – 91%). Individuals in GS and UKB samples were predominantly well-educated, with around 40% achieving post-secondary or degree qualifications, in line with the demographics of the wider UKB and GS cohorts as a consented health study. For DataLoch and GS cohorts, SIMD-based deprivation of individuals in each definition was in-line with population estimates, centred on the third quintile. For the UKB cohort, Townsend Deprivation scores indicated slightly higher area-level material deprivation (i.e., above the population median of 0) across all definitions.

For the DataLoch cohort, multinomial logistic regression, using the 2+ switch definition as reference, demonstrated that increasing age was significantly associated with slightly higher odds of being included in the 1+ switch definitions, and in the definitions that included between-class switches (ORs 1.01; see Figure 2). In contrast, increasing age was associated with slightly lower odds of being included in the 3+ switch and 3+ switch with augmentation definitions (ORs 0.99). Sex was not significantly associated with the odds of inclusion in any definition beyond the 2+ switch definition (ORs 0.91 – 1.07). Individuals from more deprived areas were more likely to be included in 1+ switch definitions (ORs around 0.95) and in definitions that included between-class switches (ORs 0.95 – 0.96). There were no significant associations between ethnicity and the odds of inclusion in each definition (ORs 0.82 – 1.24), nor any significant differences in the proportion of ethnicity groups across the definitions (X^2^ = 15.21, p = 0.995).

**Figure 2.**
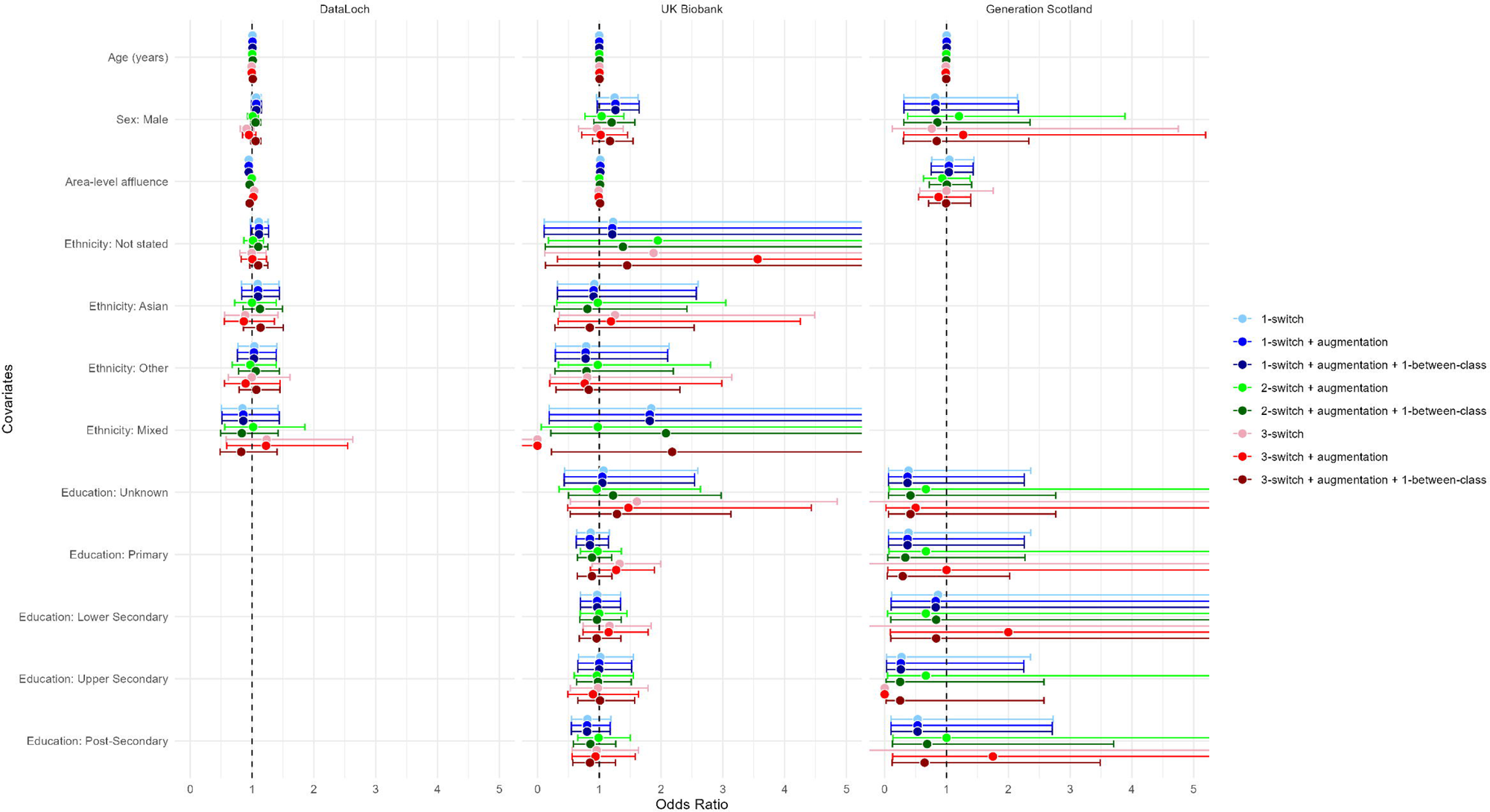
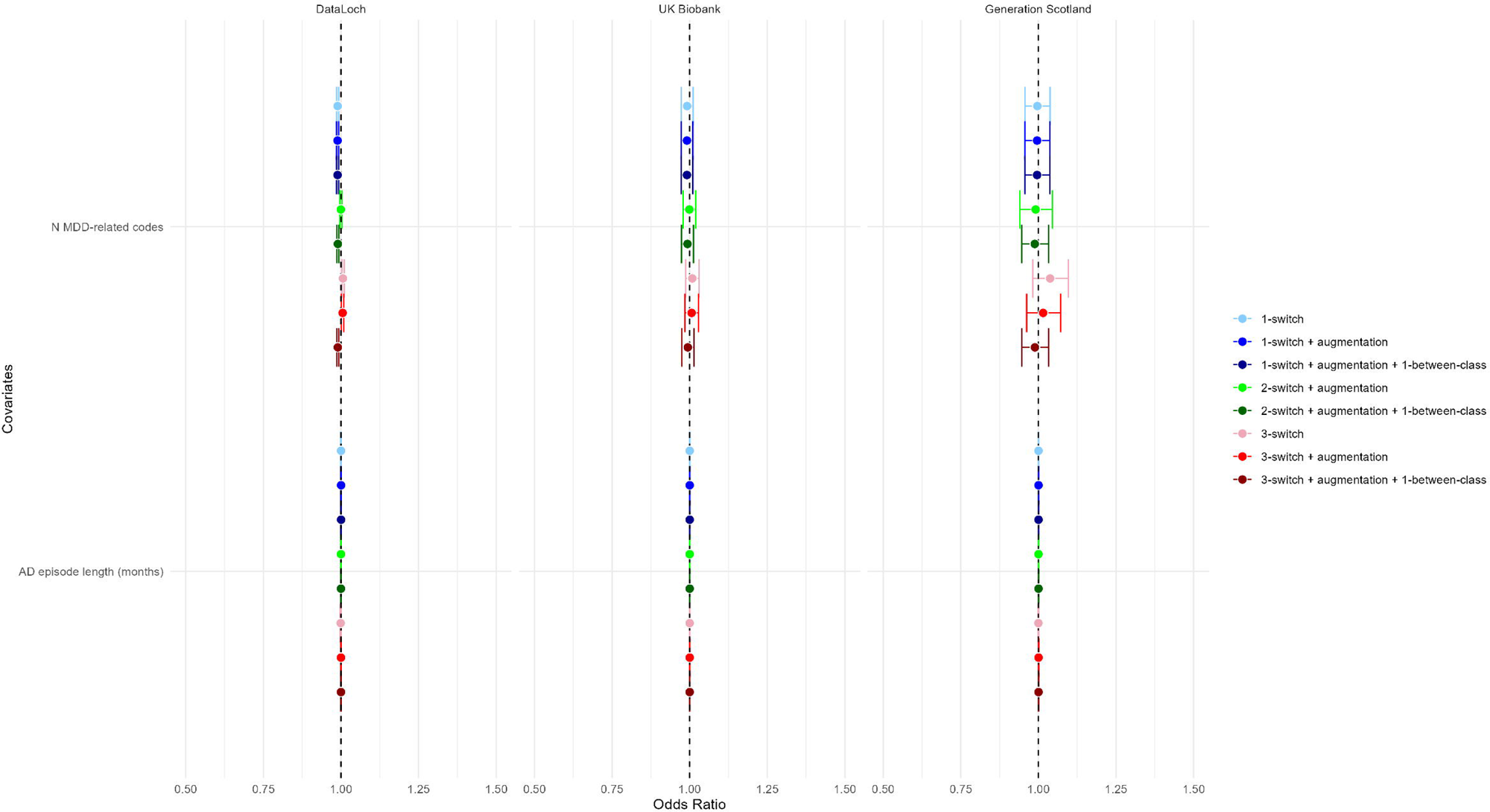
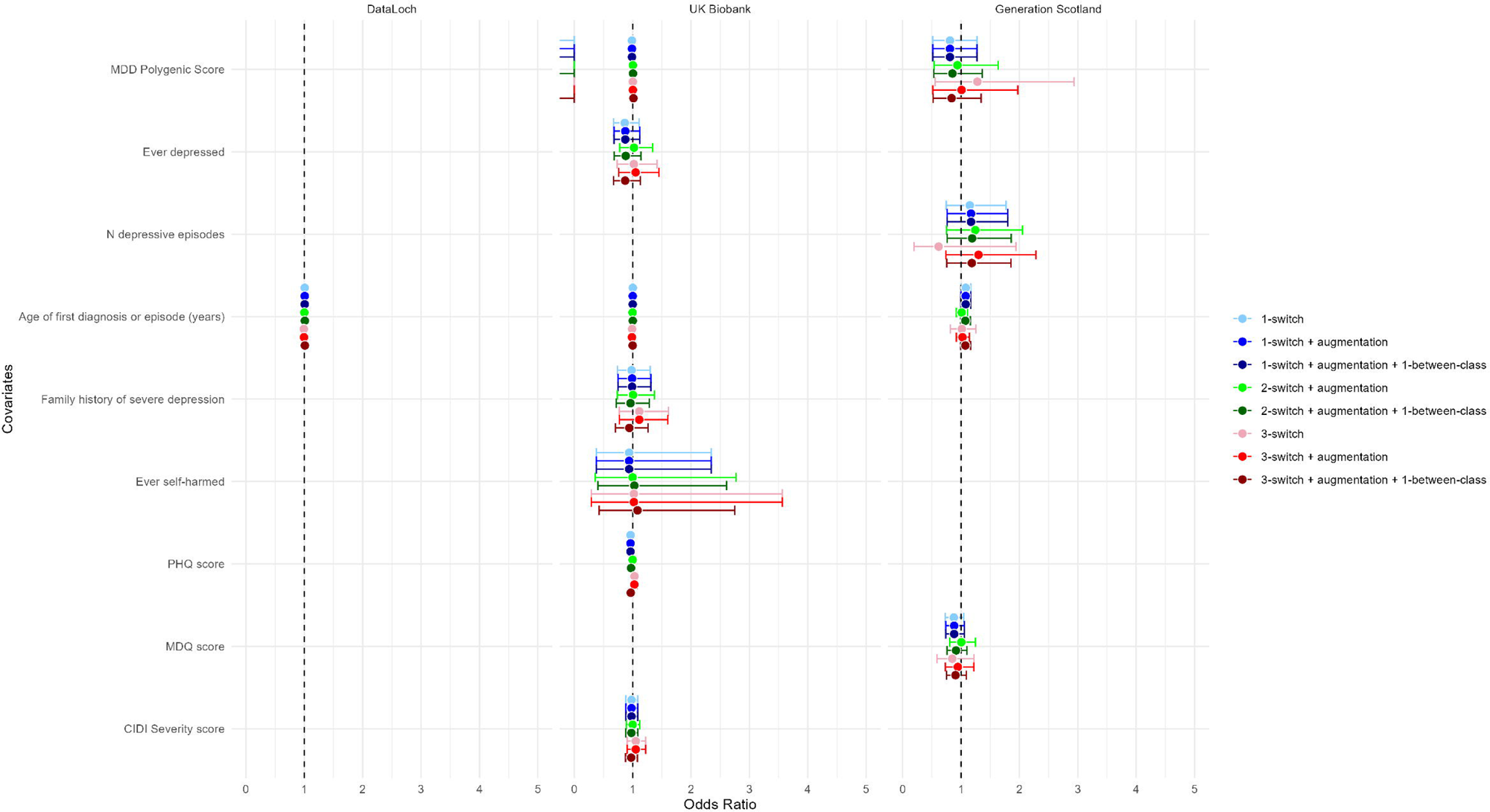
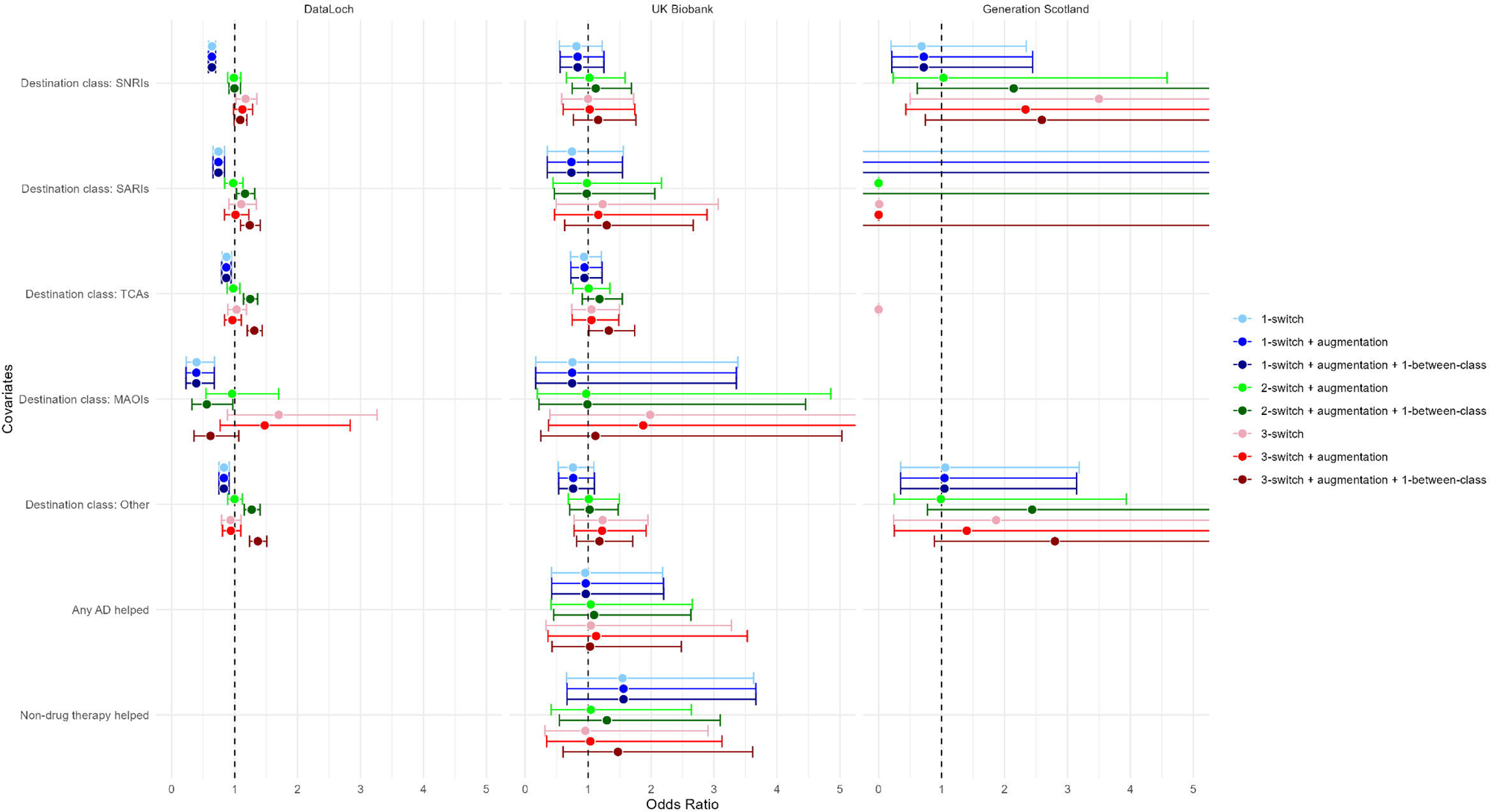
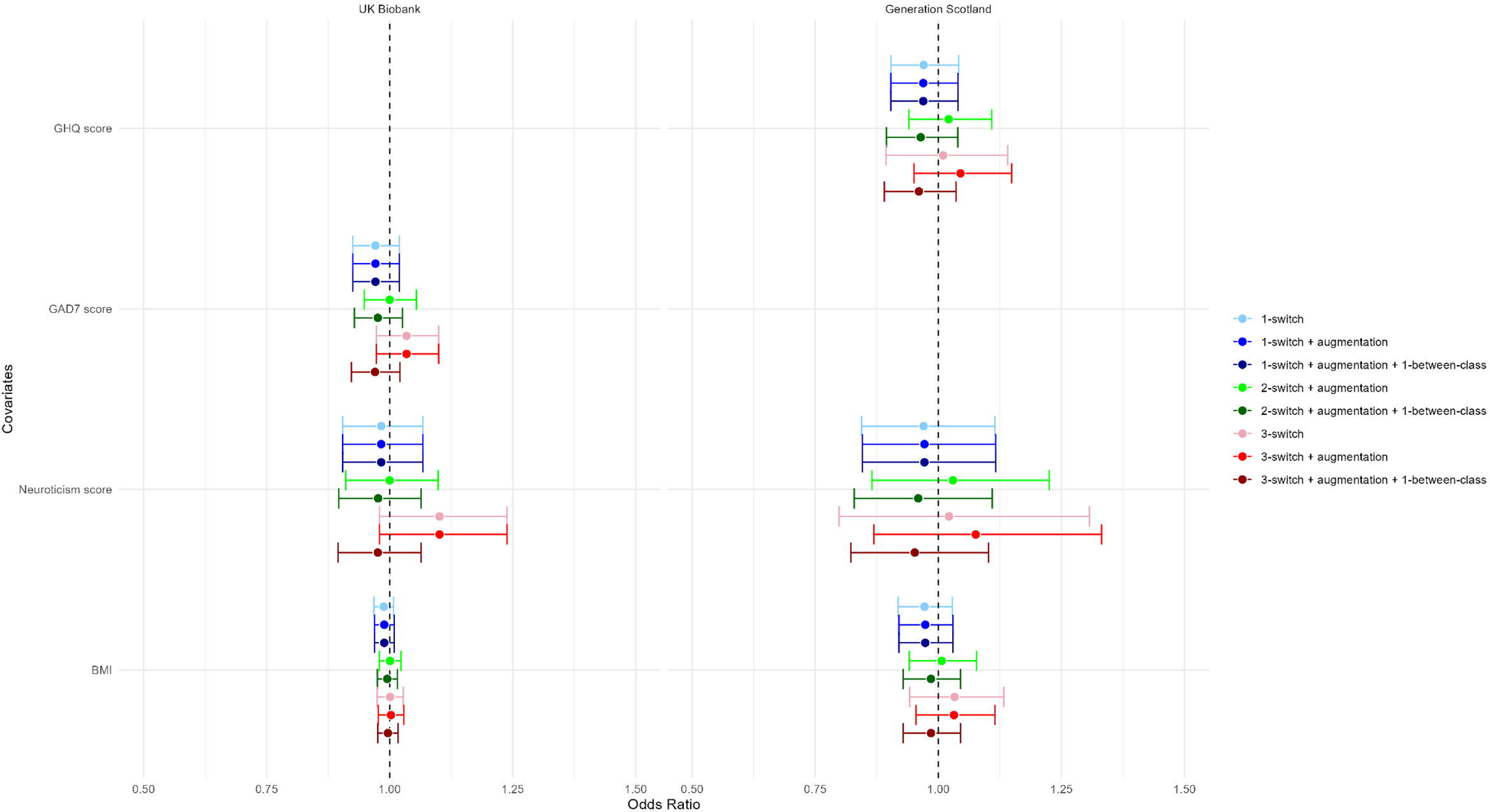
Forest plot summarising associations between covariates and the odds of being included in a Treatment Resistant Depression definition, versus the 2+ switch definition. A) Demographics, B) Clinical Presentation, C) Psychiatric Questionnaires, D) Treatment Response, E) Other covariates.

For the UKB and GS cohorts, increasing age was not significantly associated with odds of being in any definition relative to the 2+ switch definition (UKB: ORs 0.92 – 1.34; GS: ORs 0.08 – 1.80). Sex was also not significantly associated with the odds of inclusion in any given definition (UKB: ORs 0.96-1.26; GS: ORs 0.76-1.27). In terms of area-level affluence, there was no significant association between SIMD quintile and odds of being included in any of the definitions (ORs 0.75 – 1.10) nor between reversed Townsend Deprivation score and odds of being included in any of the definitions (ORs 0.99 – 1.02). In the UKB cohort, there were no significant differences in the proportion of ethnicity groups across the definitions (X^2^ = 6.60, p = 1.000).

### Clinical presentation from Electronic Health Records

The definitions based on 1+ switches had fewer MDD-related codes (median = 3) and shorter treatment time (median = 16 months) across their EHRs than those with 2+ or 3+ switches (median codes = 4; median time = 16-18 months). However, adding between-class switches resulted in fewer MDD codes and shorter treatment times for the definitions based on 2+ and 3+ switches.

Across all three cohorts, individuals with more MDD-related codes were slightly less likely to be included in the 1+ definitions than the 2+ definition (ORs 0.99 – 1.00), but slightly more likely to be included in the 3+ definitions (ORs 1.01 – 1.04) (Figure 2). Individuals with more MDD-related codes were also slightly less likely to be included in definitions that included between-class switches (ORs 0.99). Treatment time was not consistently associated with inclusion in a given definition relative to the 2+ switch definition (ORs ∼ 1.00).

### Psychiatric questionnaires and severity

In the UKB and GS cohorts, polygenic scores for MDD were similar between definitions. In DataLoch, the median age of the first recorded MDD diagnostic code was around middle-age (41-45 years), consistent with the UKB cohort where the self-reported age of first diagnosis was around middle-age (44-45 years). In contrast, in GS, median self-reported age of depression onset was around early adulthood across all definitions (20-27 years). Only around 25% of individuals in UKB self-reported a diagnosis of depression, and the median number of self-reported depressive episodes in GS was 0. Notably, depression onset and TRD classification may have occurred well before or after self-report in the surveys, since both UKB and GS had substantial periods of follow-up in electronic health records. Therefore, self-report may not align completely with episodes of depression and treatment. In UKB, self-reported self-harm was rare (around 6%) across all definitions, though this likely represents wider issues around reporting of self-harm in cohorts such as UKB (Davis et al., 2025). However, around 25% of TRD individuals reported a family history of severe depression regardless of definition. In terms of symptom severity, median PHQ4 scores from their worst depressive episode were lower in the 1+ definitions and in definitions including a between-class switch (around 3) than in other definitions (around 4). MDQ scores followed a similar pattern in the GS cohort, with lowest median scores among 1+ definitions and those including between-class switches. CIDI severity scores were generally higher in the 3+ definitions (around 7) than in other definitions (around 6), though notably not for the definition that included 3+ switches, augmentation and between-class switches.

In the DataLoch cohort, individuals with an older age of first depression diagnostic code were substantially more likely to appear in the 1+ switch definitions (ORs 1.01) and the definitions including a between-class switch (ORs 1.01) than the 2+ switch definition, but less likely to appear in the definition including 3+ switches (OR 0.99) or 3+ switches and augmentation (OR 0.99) (Figure 2). In the UKB and GS cohorts, there were no significant associations with odds of being included in any of the definitions (relative to the 2+ definition) for polygenic scores for MDD, self-reported depression, age of first depression onset, self-reported number of depressive episodes, family history of depression, or self-reported self-harm (all ORs around 1.00). Individuals with higher PHQ4 scores were less likely to be included in the 1+ switch definitions versus the 2+ switch definitions (ORs around 0.96), though they were not more likely to be included in the 3+ definitions (ORs 0.96 – 1.03).

There were no significant associations between CIDI severity scores (in UKB) and MDQ scores (in GS) and odds of being included in a given definition.

### Treatment response

SSRIs were the most common destination class across all cohorts and definitions (DataLoch: 25-35%; UKB: 38-44%; GS: 37-62%). In contrast, MAOIs were rare AD class across the three cohorts, with none of the GS participants experiencing MAOIs as their final recorded class. In general, 1+ switch definitions included a higher proportion of individuals with a destination class of SSRIs than 2+ and 3+ switch definitions. The distribution of destination AD classes was broadly similar between the definitions that only included switches and those that additionally included augmentations. In contrast, definitions which included between-class switches included a higher proportion of TCA and Other AD users across the cohorts.

In DataLoch, individuals whose final recorded AD class was not an SSRI were significantly less likely to be included in the 1+ switch definitions (ORs 0.39 – 0.87), with the lowest odds for MAOIs (ORs 0.39 – 0.40), SNRIs (ORs 0.64), SARIs (ORs 0.74), Others (ORs 0.83), and TCAs (ORs 0.87), in order. Patients whose final AD class was Others (ORs 1.27 – 1.37), TCAs (ORs 1.25 – 1.31), or SARIs (ORs 1.17 – 1.24) were significantly more likely to be included in the 2+ and 3+ TRD definitions that included between-class switches. Those ending on MAOIs (ORs 0.56) were significantly less likely to be included in the definition including 2+ switches (but not 3+ switches), augmentations and between-class switches. Individuals with SNRIs as their final class were significantly more likely to be included in the 3+ switch definition (OR 1.17).

In the GS cohort, individuals whose final recorded AD class was SARIs or TCAs were significantly more likely to be included in the other TRD definitions than the 2+ switch definition (ORs > 10), since the 2+ switch definition included no individuals whose final class was SARIs or TCAs. In UKB, only individuals with the final recorded AD class of TCAs were significantly more likely to be included and only in the TRD definition that included 3+ switches, augmentation and between-class switches (OR 1.33).

In UKB, only a small proportion of individuals (around 12%) reported that no antidepressant helped them during their depression treatment, regardless of TRD definition. Around 85% of individuals reported a non-drug treatment helping their depression. Neither reporting that an antidepressant helped nor reporting that a non-drug treatment helped were significantly associated with inclusion in any of the TRD definitions (ORs 0.95 – 1.12 and 0.76 – 1.59).

### Other covariates

In UKB, GAD7 scores were lowest (median = 2) in the 1+ switch definitions and those including between-class switches. In GS, GHQ scores showed a similar pattern, but were typically highest (medians around 6) in definitions that included augmentation (but not between-class switch). In both UKB and GS, neuroticism scores were broadly similar across TRD definitions (medians around 7). BMIs across cohorts and definitions were on average overweight (medians around 28), though for GS BMIs were higher (around 32) among the 2+ and 3+ switch definitions.

Across UKB and GS, there was no significant association with odds of inclusion in any given TRD definition for GAD7 scores (ORs 0.97-1.03), GHQ scores (ORs 0.96-1.02) and neuroticism scores (ORs 0.95-1.10). There was no significant association between BMI and odds of inclusion in any of the definitions, relative to the 2+ switch definition (ORs 0.97-1.03).

### Electroconvulsive Therapy

In supplementary analyses using the DataLoch cohort, adding receipt of ECT as a criteria for entry into the TRD definition did not substantially increase the sample size (<1%) from the simpler definition including number of switches and augmentation criteria (Supplementary Table S5). This is due to the relative rarity of ECT, but also reflects that most individuals have received antidepressant treatment appropriate to meet a TRD definition before ECT is delivered.

Associations between covariates and odds of inclusion in the ECT-inclusive TRD definitions followed the patterns of the main analyses. That is, relative to the 2+ switch definition, higher odds of entry in the 1+ switch definition that included augmentation and ECT was associated with older age (OR 1.01), higher deprivation (OR 0.95), fewer depression-related EHR codes (OR 0.99), older age of onset (OR 1.01), and fewer non-SSRI destination antidepressants (ORs 0.39 – 0.87). Individuals in the 2+ switch definition that included augmentation and ECT closely resembled those in the simpler 2+ switch definition, with no significant associations between odds of entry and any of the demographic, clinical, survey, response or other covariates (ORs ∼ 1.00). Higher odds of entry into the 3+ switch definition that included augmentation and ECT was significantly associated with younger age (OR 0.99), higher number of depression-related EHR codes (OR 1.01) and longer antidepressant treatment time (OR 1.00).

## Discussion

The present study examined how changing the TRD definition from 2+ antidepressant switches affects the resulting sample in terms of demographics, clinical characteristics, symptom severity, treatment response, and other covariates. In general, lowering the number of switches required resulted in more individuals being included in the TRD definition, and the resulting sample was older and more socioeconomically deprived, but also less severe (fewer MDD-related diagnostic codes and lower PHQ4 scores) and with older age of onset than the standard 2+ switch definition. In contrast, increasing the number of switches required resulted in fewer individuals being included in the TRD definition, and the resulting sample was younger and more female, presenting with more complex depression (more distinct MDD-related diagnostic codes) and with younger age of onset. Notably, more restrictive definitions were not associated with greater symptom severity, though statistical power was limited by small cell counts in the consented health cohorts.

Incorporating treatment information beyond switches had more mixed effects on sample size and characteristics. Adding those with an augmentation episode (i.e., switches or augmentation) resulted in a small (1%) increase in sample size in each cohort, with little change in the characteristics of the resulting sample. However, adding those with a single between-class switch (i.e., switches, augmentation or a between-class switch) substantially increased the sample size. This latter increase likely represents those individuals further down the treatment line when data began or older adults whose treatment started before widespread adoption of SSRIs, hence the older age and higher proportion of TCAs in this definition. Finally, adding those receiving ECT did not substantially increase sample size or change the resulting sample in terms of descriptives or associations with odds of entry.

Given that identifying instances of ECT requires an additional data source – and so additional time, effort and expense – the return on investment is likely low for researchers looking to classify TRD.

Notably, we also examined how these definitions changed across a cohort defined from population health records and two consented health cohorts. The most prevalent definition, based on 2+ antidepressant switches, included around 10% of MDD-positive individuals in the e-cohort created from population health records, but the rate of TRD was substantially and consistently lower (around 5%) in consented health cohorts, suggesting that TRD is underrepresented in such studies. The difference in proportions was accentuated as the number of switches required to be included increased. However, definition-related differences in demographic, clinical and treatment response characteristics were consistent across cohorts.

In the UKB cohort, TRD defined by electronic health records did not correlate well with self-reported response with most individuals, across all TRD definitions, reporting that antidepressants helped their depression. There was no evidence that self-reported responders were less likely to be included in more restrictive definitions. This may indicate sampling bias, in that individuals with less severe depression may be more likely to complete the mental health surveys. However, it may also indicate the disconnect between patients’ subjective experiences of treatment and their actual treatment, and highlights the need to contextualise routinely-collected data with patient-reported outcomes.

These results have several implications for studies of TRD, particularly for clinical trials. Firstly, prescribing/dispensing data can be combined to make nuanced definitions of TRD that better reflect real-world treatment. Adding other data sources, such as records of ECT, does not substantially increase the number of TRD patients beyond what can be captured by prescribing/dispensing data. Studies such as clinical trials that want to capture a group with more severe depression may benefit from using a TRD definition that includes 2+ switches or augmentation, as this maximises sample size and symptom severity with little change in clinical or demographic profile. While adding a between-class switch substantially increased the resulting sample size (almost double), it also resulted in an older and less severe cohort, making it a less confident compromise, particularly for clinical trials. This being said, future studies may benefit from incorporating several features (e.g., switches, augmentation and between-class switches) and multiple data sources (e.g., ECT) if available. This would allow for a continuum of TRD, similar to previous clinical staging models (e.g., Fekadu et al., 2018), and for stratification of TRD to better understand non-response.

### Limitations

The TRD definitions examined in the present study only cover the most common definitions, and do not include several features considered in other definitions. For example, we did not incorporate information on symptom severity, functional impairment or episode duration, features included in other definitions such as the Dutch Measure for quantification of Treatment Resistant Depression (Peeters et al., 2016) and the Maudsley Staging Model (Fekadu et al., 2018). These features are not easily extracted from electronic health records, as they are often not represented in the clinical codes used, and instead may reside within clinical free-text not widely accessible to researchers (Jackson et al., 2017). Furthermore, the TRD definitions included in the main analyses focussed on pharmacological therapies, and did not include non-pharmacological therapies that are sometimes included in other definitions of TRD (McIntyre et al., 2023). While many existing TRD definitions are agnostic about the kinds of treatment patients should have experienced, the reality is that the quality of data relating to these non-pharmacological therapies is poor, and many are administered in private clinics not captured by electronic health records. While supplementary analyses does include ECT, an indicator of treatment resistance when used in cases of depression, necessary data was only available in one data set, and is not available on a national scale.

Prescribing and dispensing data only approximates antidepressant exposure, as it does not track use or adherence to medication. While these can be estimated (e.g., from the ratio between the prescribed and actual frequency of collection), actual exposure can only be measured through more detailed follow-up than is available from routinely-collected health records. Longitudinal studies, particularly those with frequent biological sampling, may help to validate the TRD definitions examined here.

Although consistent in direction, there was limited statistical power to detect associations in the UKB and GS cohorts due to small sample size, particularly in less inclusive TRD definitions. Sample size was primarily affected by selecting down to those with a depression diagnosis and those with exposure antidepressant treatment. Better statistical power to examine correlates of treatment resistance may be gained from more disease-specific cohorts, such as the GLAD study (Davies et al., 2019).

## Conclusion

Understanding treatment resistance in depression requires clear definitions of TRD. However, small changes to the definition used result in substantial changes to the sample captured, both in terms of size and underlying characteristics. At the same time, definitions have a different impact depending on the population, with TRD being underrepresented in consented cohort studies. Researchers should think flexibility about treatment resistance, adapting their definition depending on the initial population, the desired sample and the available data sources. In most studies, treatment resistance can be conceived as a continuum, but for classification purposes a definition that includes 2+ switches and augmentations balances inclusivity and severity while only requiring one source of data.

## Supporting information

Clinical code list

Antidepressant Class list

Antidepressant augmentation list

## Acknowledgments

This work uses data provided by patients and collected by the NHS as part of their care and support. This project has been facilitated by the DataLoch service (reference: DL_2023_066). DataLoch enables access to de-identified extracts of health care data from the South-East Scotland region to approved applicants: dataloch.org. This research has been conducted using the UK Biobank Resource under application number 4844. The authors are grateful to all the people and their families who have taken part in GS to date, the general practitioners and the Scottish School of Primary Care for their help in recruiting them, the GS scientific steering committee and the whole GS team, which includes interviewers, computer and laboratory technicians, clerical workers, research scientists, volunteers, managers, receptionists, healthcare assistants and nurses.

## Data availability

Data are available on reasonable request from their respective data controllers after ethical and access approvals. The DataLoch data can be accessed through DataLoch (www.dataloch.org) following successful application and approvals. The UK Biobank data used in the present study is available from UK Biobank with restrictions applied. Data were used under license and thus not publicly available. Access to the UK Biobank data can be requested through a standard protocol (https://www.ukbiobank.ac.uk/register-apply/).

Researchers may request access to Generation Scotland data through their website (https://www.ed.ac.uk/generation-scotland/for-researchers).

