## Supplementary Tables S1-S6 for "Treatment Resistant Depression in electronic health records: Definitions Matter"

**Table S1.** Number of individuals removed during data cleaning due to the exclusion criteria of other psychiatric diagnoses. Note that diagnoses are not mutually exclusive.

|  | **DataLoch**  (N excluded = 26,949) | **UK Biobank**  (N excluded = 3,671) | **Generation Scotland**  (N excluded = 196) |
| --- | --- | --- | --- |
| **Drug misuse** | 10,085 | 628 | 80 |
| **Alcohol misuse** | 21,417 | 2,672 | 122 |
| **Bipolar disorder** | 1,358 | 617 | 40 |
| **Psychosis** | 1,912 | 514 | 28 |

**Table S2.** Descriptive statistics for individuals included in each definition, DataLoch cohort (N = 51,283). P-values relate to Kruskal-Wallis tests (for numeric variables) and Chi-Square tests (for categorical variables) of between-definition differences.

|  | **1+ switch (N=13,643)** | **1+ switch**  **Augmentation (N=13,729)** | **1+ switch**  **Augmentation + 1+ between-class**  **(N=13,729)** | **2+ switch (N=5,331)** | **2+ switch**  **Augmentation (N=5,495)** | **2+ switch**  **Augmentation**  **1+ between-class**  **(N=11,972)** | **3+ switch (N=2,053)** | **3+ switch**  **Augmentation (N=2,279)** | **3+ switch**  **Augmentation**  **1+ between-class**  **(N=11,811)** | **p-value** |
| --- | --- | --- | --- | --- | --- | --- | --- | --- | --- | --- |
| Age at end of follow-up (years) |  |  |  |  |  |  |  |  |  | <0.001 |
| Median  (Q1, Q3) | 63.00  (55.00, 76.00) | 63.00  (55.00, 76.00) | 63.00  (55.00, 76.00) | 62.00  (54.00, 74.00) | 62.00  (54.00, 74.00) | 64.00  (55.00, 77.00) | 61.00  (54.00, 71.00) | 61.00  (54.00, 72.00) | 64.00  (55.00, 77.00) |  |
| Sex |  |  |  |  |  |  |  |  |  | 0.021 |
| Female | 10044 (73.6%) | 10103 (73.6%) | 10103 (73.6%) | 3990 (74.8%) | 4102 (74.6%) | 8835 (73.8%) | 1572 (76.6%) | 1728 (75.8%) | 8714 (73.8%) |  |
| Male | 3599 (26.4%) | 3626 (26.4%) | 3626 (26.4%) | 1341 (25.2%) | 1393 (25.4%) | 3137 (26.2%) | 481 (23.4%) | 551 (24.2%) | 3097 (26.2%) |  |
| SIMD Quintile |  |  |  |  |  |  |  |  |  | <0.001 |
| Median  (Q1, Q3) | 3.00  (2.00, 4.00) | 3.00  (2.00, 4.00) | 3.00  (2.00, 4.00) | 3.00  (2.00, 4.00) | 3.00  (2.00, 4.00) | 3.00  (2.00, 4.00) | 3.00  (2.00, 4.00) | 3.00  (2.00, 4.00) | 3.00  (2.00, 4.00) |  |
| Ethnicity |  |  |  |  |  |  |  |  |  | 0.995 |
| Asian | 194 (1.4%) | 196 (1.4%) | 196 (1.4%) | 70 (1.3%) | 72 (1.3%) | 176 (1.5%) | 24 (1.2%) | 26 (1.1%) | 175 (1.5%) |  |
| Mixed | 45 (0.3%) | 46 (0.3%) | 46 (0.3%) | 21 (0.4%) | 22 (0.4%) | 39 (0.3%) | 10 (0.5%) | 11 (0.5%) | 38 (0.3%) |  |
| Not Stated | 934 (6.8%) | 943 (6.9%) | 943 (6.9%) | 332 (6.2%) | 347 (6.3%) | 814 (6.8%) | 127 (6.2%) | 143 (6.3%) | 802 (6.8%) |  |
| Other | 158 (1.2%) | 158 (1.2%) | 158 (1.2%) | 60 (1.1%) | 60 (1.1%) | 142 (1.2%) | 23 (1.1%) | 23 (1.0%) | 141 (1.2%) |  |
| White | 12312 (90.2%) | 12386 (90.2%) | 12386 (90.2%) | 4848 (90.9%) | 4994 (90.9%) | 10801 (90.2%) | 1869 (91.0%) | 2076 (91.1%) | 10655 (90.2%) |  |
| Number of MDD codes in EHRs |  |  |  |  |  |  |  |  |  | <0.001 |
| Median  (Q1, Q3) | 3.00  (2.00, 6.00) | 3.00  (2.00, 6.00) | 3.00  (2.00, 6.00) | 4.00  (2.00, 7.00) | 4.00  (2.00, 7.00) | 3.00  (2.00, 6.00) | 4.00  (2.00, 8.00) | 4.00  (2.00, 8.00) | 3.00  (2.00, 6.00) |  |
| Treatment time (months) |  |  |  |  |  |  |  |  |  | <0.001 |
| Median  (Q1, Q3) | 16.00  (5.03, 43.99) | 16.03  (5.03, 43.99) | 16.03  (5.03, 43.99) | 17.02  (5.98, 44.97) | 17.97  (5.98, 45.99) | 16.03  (5.03, 44.97) | 16.03  (5.98, 42.97) | 18.04  (6.05, 48.00) | 16.03  (5.03, 43.99) |  |
| Age of first MDD code (years) |  |  |  |  |  |  |  |  |  | <0.001 |
| Median  (Q1, Q3) | 45.00  (35.00, 58.00) | 45.00  (35.00, 58.00) | 45.00  (35.00, 58.00) | 43.00 (34.00, 55.00) | 43.00  (35.00, 55.00) | 45.00  (36.00, 59.00) | 41.00  (34.00, 53.00) | 42.00  (34.00, 53.00) | 46.00  (36.00, 59.00) |  |
| Destination antidepressant class |  |  |  |  |  |  |  |  |  | <0.001 |
| MAOIs | 29 (0.2%) | 29 (0.2%) | 29 (0.2%) | 24 (0.5%) | 24 (0.4%) | 27 (0.2%) | 15 (0.7%) | 15 (0.7%) | 28 (0.2%) |  |
| Other | 2179 (16.0%) | 2192 (16.0%) | 2192 (16.0%) | 859 (16.1%) | 893 (16.3%) | 2203 (18.4%) | 295 (14.4%) | 341 (15.0%) | 2224 (18.8%) |  |
| SARIs | 1039 (7.6%) | 1046 (7.6%) | 1046 (7.6%) | 458 (8.6%) | 466 (8.5%) | 1077 (9.0%) | 186 (9.1%) | 196 (8.6%) | 1075 (9.1%) |  |
| SNRIs | 2307 (16.9%) | 2321 (16.9%) | 2321 (16.9%) | 1181 (22.2%) | 1215 (22.1%) | 2376 (19.8%) | 510 (24.8%) | 560 (24.6%) | 2434 (20.6%) |  |
| SSRIs | 4776 (35.0%) | 4817 (35.1%) | 4817 (35.1%) | 1564 (29.3%) | 1629 (29.6%) | 3156 (26.4%) | 575 (28.0%) | 660 (29.0%) | 2958 (25.0%) |  |
| TCAs | 3313 (24.3%) | 3324 (24.2%) | 3324 (24.2%) | 1245 (23.4%) | 1268 (23.1%) | 3133 (26.2%) | 472 (23.0%) | 507 (22.2%) | 3092 (26.2%) |  |

**Table S3.** Descriptive statistics for individuals included in each definition, UK Biobank cohort (N = 10,556). P-values relate to Kruskal-Wallis tests (for numeric variables) and Chi-Square tests (for categorical variables) of between-definition differences.

|  | **1+ switch**  **(N=777)** | **1+ switch Augmentation**  **(N=785)** | **1+ switch Augmentation**  **1+ between-class**  **(N=785)** | **2+ switch**  **(N=475)** | **2+ switch Augmentation**  **(N=485)** | **2+ switch Augmentation**  **1+ between-class**  **(N=684)** | **3+ switch**  **(N=248)** | **3+ switch Augmentation**  **(N=260)** | **3+ switch Augmentation**  **1+ between-class**  **(N=654)** | **p-value** |
| --- | --- | --- | --- | --- | --- | --- | --- | --- | --- | --- |
| Age^1^ |  |  |  |  |  |  |  |  |  | 0.988 |
| Median  (Q1, Q3) | 56.00  (48.00, 62.00) | 56.00  (48.00, 62.00) | 56.00  (48.00, 62.00) | 56.00  (48.00, 62.00) | 56.00  (48.00, 62.00) | 56.00  (49.00, 62.00) | 56.50  (49.00, 62.00) | 57.00  (49.00, 62.00) | 57.00  (49.00, 62.00) |  |
| Sex^1^ |  |  |  |  |  |  |  |  |  | 0.455 |
| Female | 586 (75.4%) | 591 (75.3%) | 591 (75.3%) | 375 (78.9%) | 381 (78.6%) | 522 (76.3%) | 199 (80.2%) | 207 (79.6%) | 502 (76.8%) |  |
| Male | 191 (24.6%) | 194 (24.7%) | 194 (24.7%) | 100 (21.1%) | 104 (21.4%) | 162 (23.7%) | 49 (19.8%) | 53 (20.4%) | 152 (23.2%) |  |
| Townsend Deprivation^1^ |  |  |  |  |  |  |  |  |  | 0.755 |
| N-Miss | 1 | 1 | 1 | 1 | 1 | 1 | 0 | 0 | 1 |  |
| Median  (Q1, Q3) | -0.14  (-2.74, 3.71) | -0.14  (-2.74, 3.69) | -0.14  (-2.74, 3.69) | 0.04  (-2.58, 4.26) | 0.02  (-2.60, 4.17) | -0.10  (-2.74, 3.65) | 0.16  (-2.78, 4.42) | 0.16  (-2.78, 4.41) | -0.10  (-2.74, 3.59) |  |
| Educational qualification |  |  |  |  |  |  |  |  |  | 0.999 |
| N-Miss | 1 | 1 | 1 | 1 | 1 | 1 | 0 | 0 | 1 |  |
| Primary | 218 (28.1%) | 219 (27.9%) | 219 (27.9%) | 143 (30.2%) | 145 (30.0%) | 195 (28.6%) | 87 (35.1%) | 91 (35.0%) | 186 (28.5%) |  |
| Lower Secondary | 163 (21.0%) | 165 (21.0%) | 165 (21.0%) | 96 (20.3%) | 98 (20.2%) | 140 (20.5%) | 52 (21.0%) | 54 (20.8%) | 132 (20.2%) |  |
| Upper Secondary | 214 (26.5%) | 218 (26.7%) | 218 (26.7%) | 122 (24.5%) | 127 (24.9%) | 187 (26.2%) | 57 (21.6%) | 62 (22.2%) | 178 (26.1%) |  |
| Post-Secondary | 91 (11.7%) | 92 (11.7%) | 92 (11.7%) | 65 (13.7%) | 66 (13.6%) | 84 (12.3%) | 28 (11.3%) | 29 (11.2%) | 79 (12.1%) |  |
| Degree | 208 (26.8%) | 212 (27.0%) | 212 (27.0%) | 117 (24.7%) | 122 (25.2%) | 181 (26.5%) | 55 (22.2%) | 60 (23.1%) | 174 (26.6%) |  |
| Unknown | 15 (1.9%) | 15 (1.9%) | 15 (1.9%) | 8 (1.7%) | 8 (1.7%) | 15 (2.2%) | 6 (2.4%) | 6 (2.3%) | 15 (2.3%) |  |
| Ethnicity^1^ |  |  |  |  |  |  |  |  |  | 1.000 |
| N-Miss | 3 | 3 | 3 | 3 | 3 | 3 | 1 | 1 | 2 |  |
| Asian | 9 (1.2%) | 9 (1.2%) | 9 (1.2%) | 6 (1.3%) | 6 (1.2%) | 7 (1.0%) | 4 (1.6%) | 4 (1.5%) | 7 (1.1%) |  |
| Mixed | 3 (0.4%) | 3 (0.4%) | 3 (0.4%) | 1 (0.2%) | 1 (0.2%) | 3 (0.4%) | 0 (0.0%) | 0 (0.0%) | 3 (0.5%) |  |
| Not Stated | 2 (0.3%) | 2 (0.3%) | 2 (0.3%) | 1 (0.2%) | 2 (0.4%) | 2 (0.3%) | 1 (0.4%) | 2 (0.8%) | 2 (0.3%) |  |
| Other | 9 (1.2%) | 9 (1.2%) | 9 (1.2%) | 7 (1.5%) | 7 (1.5%) | 8 (1.2%) | 3 (1.2%) | 3 (1.2%) | 8 (1.2%) |  |
| White | 751 (97.0%) | 759 (97.1%) | 759 (97.1%) | 457 (96.8%) | 466 (96.7%) | 661 (97.1%) | 239 (96.8%) | 250 (96.5%) | 632 (96.9%) |  |
| Number of MDD codes in EHRs |  |  |  |  |  |  |  |  |  | 0.958 |
| Median  (Q1, Q3) | 1.00 (1.00, 3.00) | 1.00  (1.00, 3.00) | 1.00  (1.00, 3.00) | 2.00  (1.00, 3.00) | 2.00  (1.00, 3.00) | 1.00  (1.00, 3.00) | 2.00  (1.00, 3.00) | 2.00  (1.00, 3.00) | 1.00  (1.00, 3.00) |  |
| Treatment time (months) |  |  |  |  |  |  |  |  |  | 0.522 |
| Median  (Q1, Q3) | 5.91  (0.00, 27.10) | 6.08  (0.00, 27.43) | 6.08  (0.00, 27.43) | 5.98  (0.00, 27.61) | 6.34  (0.00, 29.70) | 5.39  (0.00, 23.80) | 5.63  (0.00, 27.34) | 6.47  (0.00, 32.00) | 4.44  (0.00, 22.31) |  |
| MDD Polygenic Risk Score |  |  |  |  |  |  |  |  |  | 1.000 |
| N-Miss | 27 | 28 | 28 | 21 | 22 | 27 | 9 | 10 | 25 |  |
| Median (Q1, Q3) | 0.00  (0.00, 0.00) | 0.00  (0.00, 0.00) | 0.00  (0.00, 0.00) | 0.00  (0.00, 0.00) | 0.00  (0.00, 0.00) | 0.00  (0.00) | 0.00 (0.00) | 0.00  (0.00) | 0.00  (0.00) |  |
| Self-reported depression^2^ |  |  |  |  |  |  |  |  |  | 0.715 |
| N-Miss | 1 | 1 | 1 | 1 | 1 | 1 | 0 | 0 | 1 |  |
| No | 584 (75.3%) | 588 (75.0%) | 588 (75.0%) | 344 (72.6%) | 348 (71.9%) | 513 (75.1%) | 179 (72.2%) | 185 (71.2%) | 491 (75.2%) |  |
| Yes | 192 (24.7%) | 196 (25.0%) | 196 (25.0%) | 130 (27.4%) | 136 (28.1%) | 170 (24.9%) | 69 (27.8%) | 75 (28.8%) | 162 (24.8%) |  |
| Self-reported age of first diagnosis^3^ |  |  |  |  |  |  |  |  |  | 0.913 |
| N-Miss | 585 | 589 | 589 | 345 | 349 | 514 | 179 | 185 | 492 |  |
| Median  (Q1, Q3) | 45.52  (35.40, 53.74) | 45.25  (35.23, 53.74) | 45.25  (35.23, 53.74) | 44.54  (33.47, 53.11) | 44.48  (32.43, 52.90) | 45.25  (34.81, 53.99) | 44.46  (30.50, 51.59) | 44.28  (30.39, 51.54) | 44.92  (34.72, 53.99) |  |
| Family history of severe depression^2^ |  |  |  |  |  |  |  |  |  | 0.989 |
| N-Miss | 121 | 122 | 122 | 84 | 86 | 114 | 42 | 45 | 110 |  |
| No | 497 (75.8%) | 501 (75.6%) | 501 (75.6%) | 295 (75.4%) | 300 (75.2%) | 434 (76.1%) | 151 (73.3%) | 157 (73.0%) | 416 (76.5%) |  |
| Yes | 159 (24.2%) | 162 (24.4%) | 162 (24.4%) | 96 (24.6%) | 99 (24.8%) | 136 (23.9%) | 55 (26.7%) | 58 (27.0%) | 128 (23.5%) |  |
| Self-reported self harm^3^ |  |  |  |  |  |  |  |  |  | 1 |
| N-Miss | 603 | 611 | 611 | 374 | 384 | 537 | 200 | 212 | 514 |  |
| No | 163 (93.7%) | 163 (93.7%) | 163 (93.7%) | 94 (93.1%) | 94 (93.1%) | 137 (93.2%) | 45 (93.8%) | 45 (93.8%) | 130 (92.9%) |  |
| Yes | 11 (6.3%) | 11 (6.3%) | 11 (6.3%) | 7 (6.9%) | 7 (6.9%) | 10 (6.8%) | 3 (6.2%) | 3 (6.2%) | 10 (7.1%) |  |
| PHQ4 score^3^ |  |  |  |  |  |  |  |  |  | <0.001 |
| N-Miss | 17 | 17 | 17 | 12 | 12 | 15 | 7 | 7 | 15 |  |
| Median  (Q1, Q3) | 3.00  (1.00, 5.00) | 3.00  (1.00, 5.00) | 3.00  (1.00, 5.00) | 4.00  (1.00, 5.00) | 4.00  (1.00, 5.00) | 3.00  (1.00, 5.00) | 4.00  (2.00, 6.00) | 4.00  (1.00, 6.00) | 3.00  (1.00, 5.00) |  |
| CIDI severity^2^ |  |  |  |  |  |  |  |  |  | 0.805 |
| N-Miss | 602 | 610 | 610 | 373 | 383 | 536 | 199 | 211 | 513 |  |
| Median  (Q1, Q3) | 6.00  (5.00, 7.50) | 6.00  (5.00, 7.50) | 6.00  (5.00, 7.50) | 6.00  (5.00, 8.00) | 6.00  (5.00, 8.00) | 6.00  (5.00, 7.00) | 7.00  (5.00, 8.00) | 7.00  (5.00, 8.00) | 6.00  (5.00, 7.00) |  |
| Destination antidepressant class |  |  |  |  |  |  |  |  |  | 0.692 |
| MAOIs | 4 (0.5%) | 4 (0.5%) | 4 (0.5%) | 3 (0.6%) | 3 (0.6%) | 4 (0.6%) | 3 (1.2%) | 3 (1.2%) | 4 (0.6%) |  |
| Other | 78 (10.0%) | 79 (10.1%) | 79 (10.1%) | 58 (12.2%) | 60 (12.4%) | 79 (11.5%) | 35 (14.1%) | 37 (14.2%) | 82 (12.5%) |  |
| SARIs | 16 (2.1%) | 16 (2.0%) | 16 (2.0%) | 12 (2.5%) | 12 (2.5%) | 16 (2.3%) | 7 (2.8%) | 7 (2.8%) | 19 (2.9%) |  |
| SNRIs | 66 (8.5%) | 68 (8.7%) | 68 (8.7%) | 47 (9.9%) | 49 (10.1%) | 70 (10.2%) | 23 (9.3%) | 25 (9.6%) | 63 (9.6%) |  |
| SSRIs | 345 (44.4%) | 346 (44.1%) | 346 (44.1%) | 194 (40.8%) | 195 (40.2%) | 258 (37.7%) | 96 (38.7%) | 99 (38.1%) | 230 (35.2%) |  |
| TCAs | 268 (34.5%) | 272 (34.6%) | 272 (34.6%) | 161 (33.9%) | 166 (34.2%) | 257 (37.6%) | 84 (33.9%) | 89 (34.2%) | 256 (39.1%) |  |
| Any antidepressant helped^3^ |  |  |  |  |  |  |  |  |  | 1 |
| N-Miss | 664 | 651 | 651 | 398 | 406 | 577 | 210 | 220 | 551 |  |
| No | 18 (13.5%) | 18 (13.4%) | 18 (13.4%) | 10 (13.0%) | 10 (12.7%) | 13 (12.1%) | 5 (11.9%) | 5 (11.1%) | 13 (12.1%) |  |
| Yes | 115 (86.5%) | 116 (86.6%) | 116 (86.6%) | 67 (87.0%) | 69 (87.3%) | 94 (87.9%) | 33 (86.8%) | 35 (87.5%) | 88 (87.1%) |  |
| Non-drug treatment helped^3^ |  |  |  |  |  |  |  |  |  | 0.989 |
| N-Miss | 665 | 672 | 672 | 418 | 426 | 594 | 218 | 228 | 568 |  |
| No | 15 (12.7%) | 15 (12.6%) | 15 (12.6%) | 11 (18.3%) | 11 (17.5%) | 14 (14.9%) | 6 (18.2%) | 6 (16.7%) | 12 (13.3%) |  |
| Yes, at least a little | 103 (87.3%) | 104 (87.4%) | 104 (87.4%) | 49 (81.7%) | 52 (82.5%) | 80 (85.1%) | 27 (81.8%) | 30 (83.3%) | 78 (86.7%) |  |
| GAD7 severity^3^ |  |  |  |  |  |  |  |  |  | 0.130 |
| N-Miss | 602 | 610 | 610 | 373 | 383 | 536 | 199 | 211 | 513 |  |
| Median  (Q1, Q3) | 2.00  (0.00, 6.00) | 2.00  (0.00, 6.00) | 2.00  (0.00, 6.00) | 4.00  (0.00, 6.75) | 4.00  (0.00, 6.75) | 2.00  (0.00, 6.00) | 5.00  (0.00, 8.00) | 5.00  (0.00, 8.00) | 2.00  (0.00, 6.00) |  |
| Neuroticism score^2^ |  |  |  |  |  |  |  |  |  | 0.279 |
| N-Miss | 633 | 641 | 641 | 394 | 404 | 564 | 207 | 219 | 539 |  |
| Median  (Q1, Q3) | 7.00  (4.00, 9.00) | 7.00  (4.00, 9.00) | 7.00  (4.00, 9.00) | 7.00  (4.00, 9.00) | 7.00  (4.00, 9.00) | 6.50  (3.75, 9.00) | 8.00  (5.00, 10.00) | 8.00  (5.00, 10.00) | 6.00  (3.50, 9.00) |  |
| BMI |  |  |  |  |  |  |  |  |  | 0.852 |
| N-Miss | 3 | 4 | 4 | 2 | 3 | 4 | 0 | 1 | 4 |  |
| Median  (Q1,Q3) | 27.88  (24.87, 31.11) | 27.91  (24.87, 31.14) | 27.91  (24.87, 31.14) | 28.20  (25.01, 31.94) | 28.36  (25.02, 32.06) | 28.16  (25.01, 31.56) | 28.27  (25.03, 31.83) | 28.40  (25.03, 32.15) | 28.18  (25.02, 31.78) |  |

1. Baseline; 2. In-person assessment; 3. Online Mental Health Questionnaire

**Table S4.** Descriptive statistics for individuals included in each definition, Generation Scotland cohort (N = 649). P-values relate to Kruskal-Wallis tests (for numeric variables) and Chi-Square tests (for categorical variables) of between-definition differences.

|  | **1+ switch (N=133)** | **1+ switch Augmentation (N=136)** | **1+ switch Augmentation**  **1+ between-class (N=136)** | **2+ switch (N=23)** | **2+ switch Augmentation (N=29)** | **2+ switch Augmentation**  **1+ between-class (N=81)** | **3+ switch (N=8)** | **3+ switch**  **Augmentation (N=14)** | **3+ switch Augmentation**  **1+ between-class (N=78)** | **p-value** |
| --- | --- | --- | --- | --- | --- | --- | --- | --- | --- | --- |
| Age at survey (years) |  |  |  |  |  |  |  |  |  | 0.963 |
| Median  (Q1, Q3) | 46.00  (36.00, 59.00) | 46.50  (36.00, 59.00) | 46.50  (36.00, 59.00) | 45.00  (35.50, 52.50) | 43.00  (35.00, 53.00) | 43.00  (33.00, 58.00) | 42.00  (39.00, 49.75) | 40.50  (36.25, 51.25) | 43.00  (33.50, 58.75) |  |
| Sex |  |  |  |  |  |  |  |  |  | 0.992 |
| Female | 98 (73.7%) | 100 (73.5%) | 100 (73.5%) | 16 (69.6%) | 19 (65.5%) | 59 (72.8%) | 6 (75.0%) | 9 (64.3%) | 57 (73.1%) |  |
| Male | 35 (26.3%) | 36 (26.5%) | 36 (26.5%) | 7 (30.4%) | 10 (34.5%) | 22 (27.2%) | 2 (25.0%) | 5 (35.7%) | 21 (26.9%) |  |
| SIMD Quintile |  |  |  |  |  |  |  |  |  | 0.988 |
| N-Miss | 10 | 10 | 10 | 2 | 2 | 7 | 0 | 0 | 7 |  |
| Median  (Q1, Q3) | 3.00  (2.00, 4.00) | 3.00  (2.00, 4.00) | 3.00  (2.00, 4.00) | 3.00  (1.00, 4.00) | 3.00  (1.00, 4.00) | 3.00  (2.00, 4.00) | 3.50  (1.00, 4.25) | 2.50  (1.25, 4.00) | 3.00  (2.00, 4.00) |  |
| Educational qualification |  |  |  |  |  |  |  |  |  | 1.000 |
| N-Miss | 5 | 5 | 5 | 1 | 2 | 5 | 0 | 1 | 5 |  |
| Primary | 17 (13.3%) | 17 (13.0%) | 17 (13.0%) | 4 (18.2%) | 4 (14.8%) | 8 (10.5%) | 2 (25.0%) | 2 (15.4%) | 7 (9.6%) |  |
| Lower Secondary | 19 (14.8%) | 19 (14.5%) | 19 (14.5%) | 2 (9.1%) | 2 (7.4%) | 10 (13.2%) | 2 (25.0%) | 2 (15.4%) | 10 (13.7%) |  |
| Upper Secondary | 6 (4.7%) | 6 (4.6%) | 6 (4.6%) | 2 (9.1%) | 2 (7.4%) | 3 (3.9%) | 0 (0.0%) | 0 (0.0%) | 3 (4.1%) |  |
| Post-Secondary | 47 (36.7%) | 49 (37.4%) | 49 (37.4%) | 8 (36.4%) | 12 (44.4%) | 33 (43.4%) | 3 (37.5%) | 7 (53.8%) | 31 (42.5%) |  |
| Degree | 22 (17.2%) | 23 (17.6%) | 23 (17.6%) | 2 (9.1%) | 3 (11.1%) | 12 (15.8%) | 0 (0.0%) | 1 (7.7%) | 12 (16.4%) |  |
| Unknown | 17 (13.3%) | 17 (13.0%) | 17 (13.0%) | 4 (18.2%) | 4 (14.8%) | 10 (13.2%) | 1 (12.5%) | 1 (7.7%) | 10 (13.7%) |  |
| Number of MDD codes in EHRs |  |  |  |  |  |  |  |  |  | 0.990 |
| Median  (Q1, Q3) | 3.00  (2.00, 6.00) | 3.00  (2.00, 5.25) | 3.00  (2.00, 5.25) | 3.00  (2.00, 5.50) | 3.00  (2.00, 5.00) | 3.00  (2.00, 5.00) | 5.50  (2.00, 27.50) | 3.50  (2.00, 5.75) | 3.00  (2.00, 5.00) |  |
| Treatment time (months) |  |  |  |  |  |  |  |  |  | 0.861 |
| Median  (Q1, Q3) | 17.94  (4.99, 48.00) | 18.51  (4.99, 50.00) | 18.51  (4.99, 50.00) | 10.02  (3.50, 27.97) | 20.99  (4.96, 39.03) | 22.01  (4.96, 52.96) | 12.98  (2.77, 27.47) | 23.98  (5.98, 62.02) | 23.51  (4.98, 52.72) |  |
| MDD Polygenic Risk Score |  |  |  |  |  |  |  |  |  | 0.838 |
| N-Miss | 7 | 7 | 7 | 1 | 1 | 3 | 0 | 0 | 3 |  |
| Median  (Q1, Q3) | 0.35  (-0.20, 1.14) | 0.35  (-0.22, 1.14) | 0.35  (-0.22, 1.14) | 0.77  (0.20, 1.44) | 0.70  (-0.17, 1.43) | 0.39  (-0.20, 1.28) | 1.43  (0.03, 1.55) | 1.02  (-0.66, 1.52) | 0.37  (-0.21, 1.24) |  |
| N self-reported depression episodes |  |  |  |  |  |  |  |  |  | 0.941 |
| N-Miss | 15 | 16 | 16 | 3 | 4 | 11 | 0 | 1 | 11 |  |
| Median  (Q1, Q3) | 0.00  (0.00, 1.00) | 0.00  (0.00, 1.00) | 0.00  (0.00, 1.00) | 0.00  (0.00, 1.25) | 0.00  (0.00, 2.00) | 0.00  (0.00, 1.00) | 0.00  (0.00, 0.00) | 0.00  (0.00, 2.00) | 0.00  (0.00, 1.00) |  |
| Self-reported age of onset |  |  |  |  |  |  |  |  |  | 0.220 |
| N-Miss | 81 | 81 | 81 | 15 | 16 | 45 | 7 | 8 | 43 |  |
| Median  (Q1, Q3) | 27.50 (19.75, 37.25) | 27.00  (19.00, 36.50) | 27.00  (19.00, 36.50) | 20.00  (17.50, 25.00) | 21.00  (17.00, 25.00) | 24.00  (17.75, 35.75) | 21.00  (21.00, 21.00) | 21.00  (18.00, 23.25) | 24.00  (18.00, 36.50) |  |
| Mood Disorder Questionnaire total score |  |  |  |  |  |  |  |  |  | 0.831 |
| N-Miss | 70 | 70 | 70 | 14 | 16 | 43 | 5 | 7 | 41 |  |
| Median  (Q1, Q3) | 3.00  (1.00, 7.00) | 3.00  (1.00, 7.00) | 3.00  (1.00, 7.00) | 6.00  (3.00, 10.00) | 6.00  (3.00, 10.00) | 4.50  (1.00, 7.00) | 0.00  (0.00, 5.50) | 6.00  (0.00, 9.00) | 4.00  (1.00, 7.00) |  |
| Destination antidepressant class |  |  |  |  |  |  |  |  |  | 0.234 |
| MAOIs | 0 (0.0%) | 0 (0.0%) | 0 (0.0%) | 0 (0.0%) | 0 (0.0%) | 0 (0.0%) | 0 (0.0%) | 0 (0.0%) | 0 (0.0%) |  |
| Other | 31 (23.3%) | 31 (22.8%) | 31 (22.8%) | 5 (21.7%) | 6 (20.7%) | 27 (33.3%) | 2 (25.0%) | 3 (21.4%) | 27 (34.6%) |  |
| SARIs | 1 (0.8%) | 1 (0.7%) | 1 (0.7%) | 0 (0.0%) | 0 (0.0%) | 1 (1.2%) | 0 (0.0%) | 0 (0.0%) | 1 (1.3%) |  |
| SNRIs | 16 (12.0%) | 17 (12.5%) | 17 (12.5%) | 4 (17.4%) | 5 (17.2%) | 19 (23.5%) | 3 (37.5%) | 4 (28.6%) | 20 (25.6%) |  |
| SSRIs | 82 (61.7%) | 83 (61.0%) | 83 (61.0%) | 14 (60.9%) | 17 (58.6%) | 31 (38.3%) | 3 (37.5%) | 6 (42.9%) | 27 (34.6%) |  |
| TCAs | 3 (2.3%) | 4 (2.9%) | 4 (2.9%) | 0 (0.0%) | 1 (3.4%) | 3 (3.7%) | 0 (0.0%) | 1 (7.1%) | 3 (3.8%) |  |
| General Health Questionnaire total score |  |  |  |  |  |  |  |  |  | 0.077 |
| N-Miss | 7 | 7 | 7 | 0 | 0 | 4 | 0 | 0 | 4 |  |
| Median  (Q1, Q3) | 2.00  (0.00, 9.00) | 2.00  (0.00, 9.00) | 2.00  (0.00, 9.00) | 4.00  (1.50, 10.50) | 6.00  (2.00, 11.00) | 2.00  (0.00, 9.00) | 4.50  (1.00, 10.25) | 7.00  (1.50, 10.75) | 2.00  (0.00, 7.75) |  |
| Neuroticism total score |  |  |  |  |  |  |  |  |  | 0.936 |
| N-Miss | 12 | 12 | 12 | 2 | 3 | 7 | 0 | 1 | 7 |  |
| Median  (Q1, Q3) | 7.00  (5.00, 9.00) | 7.00  (4.75, 9.25) | 7.00  (4.75, 9.25) | 7.00  (5.00, 9.00) | 7.00  (5.25, 9.00) | 7.00  (4.00, 9.00) | 7.50  (5.75, 9.00) | 8.00  (6.00, 9.00) | 7.00  (4.00, 9.00) |  |
| BMI |  |  |  |  |  |  |  |  |  | 0.245 |
| N-Miss | 5 | 5 | 5 | 0 | 0 | 3 | 0 | 0 | 3 |  |
| Median  (Q1, Q3) | 28.20  (24.63, 32.10) | 28.24  (24.64, 32.13) | 28.24  (24.64, 32.13) | 31.93  (24.64, 32.79) | 31.93  (25.75, 34.06) | 28.44  (24.30, 32.79) | 32.66  (29.65, 36.33) | 32.66  (28.04, 36.15) | 28.63  (24.13, 32.79) |  |

**Table S5.** Descriptive statistics for the Treatment Resistant Depression definitions that included ECT in the DataLoch cohort (N = 51,283). Column headings describe the inclusion criteria for each definition, any of which can be met for inclusion. Note descriptive statistics for the 1+ switch, augmentation and ECT definition have been censored due to small differences in sample size with the main analyses, to avoid statistical disclosure.

|  | **1+ switch**  **Augmentation**  **ECT**  **(N~13,643) (27%)** | **2+ switch**  **Augmentation**  **ECT**  **(N=5,506) (11%)** | **3+ switch**  **Augmentation**  **ECT**  **(N=2,295) (4%)** | **p-value** |
| --- | --- | --- | --- | --- |
| Age at end of follow-up (years) |  |  |  | 0.003 |
| Median  (Q1, Q3) | - | 62.00  (54.00, 74.00) | 61.00  (54.00, 72.00) |  |
| Sex |  |  |  | 0.269 |
| Female | - | 4,109 (74.6%) | 1,740 (75.8%) |  |
| Male | - | 1,397 (25.4%) | 555 (24.2%) |  |
| SIMD Quintile |  |  |  | 0.236 |
| Median  (Q1, Q3) | - | 3.00  (2.00, 4.00) | 3.00  (2.00, 4.00) |  |
| Ethnicity |  |  |  | 0.950 |
| Asian | - | 72 (1.3%) | 26 (1.1%) |  |
| Mixed | - | 22 (0.3%) | 11 (0.5%) |  |
| Not Stated | - | 348 (6.8%) | 144 (6.3%) |  |
| Other | - | 61 (1.1%) | 24 (1.0%) |  |
| White | - | 5,003 (90.9%) | 2,090 (91.1%) |  |
| Number of MDD codes in EHRs |  |  |  | <0.001 |
| Median  (Q1, Q3) | - | 4.00  (2.00, 7.00) | 4.00  (2.00, 8.00) |  |
| Treatment time (months) |  |  |  | 0.166 |
| Median  (Q1, Q3) | - | 17.97  (5.98, 45.99) | 18.04  (6.05, 48.00) |  |
| Age of first MDD code (years) |  |  |  | 0.001 |
| Median  (Q1, Q3) | - | 43.00  (35.00, 74.00) | 42.00  (34.00, 53.00) |  |
| Destination antidepressant class |  |  |  | 0.120 |
| MAOIs | - | 24 (0.4%) | 15 (0.7%) |  |
| Other | - | 896 (16.3%) | 344 (15.0%) |  |
| SARIs | - | 466 (8.5%) | 196 (8.5%) |  |
| SNRIs | - | 1,221 (22.2%) | 568 (24.7%) |  |
| SSRIs | - | 1,631 (29.6%) | 664 (28.9%) |  |
| TCAs | - | 1,268 (23.0%) | 508 (22.1%) |  |

**Table S6.** UK Biobank field codes used to identify covariates.

| **Field ID** | **Covariate** |
| --- | --- |
| f.21022 | Age |
| f.31 | Sex |
| f.22189 | Townsend deprivation index |
| f.6138 | Educational qualification |
| f.21000 | Ethnicity |
| f.29000 | Self-report depression diagnosis |
| f.29034 | Self-report age of first diagnosis |
| f.20107, f.20110, f.20111 | Family history of severe depression |
| f.29111 | Self-reported self harm |
| f.2050, f.2060, f.2070, f.2080, f.20507, f.20508, f.20510, f.20511, f.20513, f.20514, f.20517, f.20518, f.20519 | PHQ4 score |
| f.20441, f.20446, f.20441, f.20449, f.20450, f.20439, f.20440 | CIDI severity |
| f.29040, f.29041, f.29042, f.29043, f.29044, f.29045, f.29046 | Any antidepressant helped |
| f.29047, f.29048 | Non-drug treatment helped |
| f.29058, f.29059, f.29060, f.29061, f.29062, f.29063, f.29064 | GAD7 severity |
| f.20127 | Neuroticism score |
| f.21001 | BMI |
